# Treatment pathways for patients with uncontrolled diabetes and probable diabetes distress

**DOI:** 10.64898/2026.09.16.26363258

**Authors:** David A. Dorr, Katherine D. Peak, Emma Young, LeAnn Michaels, Ryan Tweet, Danielle Hessler Jones

## Abstract

Diabetes distress (DD) has a major impact on both diabetes control and emotional well-being but is mis- and underdiagnosed and undertreated. Treatment often starts with behavioral health (BH) through counseling or medications; in some patients, Certified Diabetes Care and Education Specialists (CDCES) are proven to better manage treatment burden and address distress. We performed an observational study comparing outcomes for people with high emotional distress (PHQ9 or GAD7 > 10) and high A1c 8.0 at baseline with CDCES support, BH treatment, both or neither. Results demonstrate both CDCES and behavioral health treatment were most effective (A1c −1.2%, PHQ −3.1) but received in only 4.3% of patients, while 50% of patients received BH treatment alone. Conclusion is that these services remain valuable in routine practice but are underutilized. Workflows that assist in CDCES treatment for patients at risk could improve outcomes.

## Introduction

Diabetes distress (DD) is a distinct and important behavioral health need that interferes with a person’s ability to manage their diabetes and is often mistaken for depression. DD refers to common feelings of worry, frustration, and exhaustion from managing diabetes care that impacts 45.4% of people with diabetes.^1^ Accurate identification of DD and tailored treatment mitigate risks of poor diabetes control.^2^

The American Diabetes Association (ADA) and American Psychological Association (APA) recommend approaches for patients with co-occurring diabetes and mood disorder that have strong evidence: Certified Diabetes Care and Education Specialists (CDCES) improve diabetes control (ADA recommendation) and can reduce DD.^3^ Patients with more pronounced depressive symptoms may be helped by early pharmacologic approaches to increase executive function required for diabetes self-management.^4^ Systematic reviews of co-morbid depression and emotional factors in patients with DM emphasize screening for risk factors^5^ and demonstrate effectiveness of treatment with psychotropic medications, group therapy, psycho-therapy and collaborative care to improve diabetes control and mood symptoms; similarly, fewer than half receive integrated mood treatments.^6,7^ Studies that measure DD demonstrate improvements in emotional burden and associations with improved diabetes self-management and treatment adherence.^8,9^

The APA recommends integration of general medical conditions with behavioral health (BH) to determine best treatment approaches and collaboration between the care team.^10^ Addressing psychosocial care within integrated, collaborative, patient-centered diabetes care was recommended by the ADA in 2016^9^ and re-emphasized in the 2026 Standards of Care in Diabetes.^3^ Despite recommendations, most patients do not receive evidence-based care.

## Methods

We conducted a preliminary analysis selecting patients with uncontrolled Type 2 diabetes mellitus (DM) and mood disturbance.

Using a dataset available to researchers from our institutes electronic health records, we queried adult patients seen at an ambulatory or telehealth visit from 1/2023 to 12/2025.

Uncontrolled Type 2 diabetes mellitus (DM) was defined by hemoglobin A1c (HbA1c) values, a measure of diabetes control, above 8.0% or an escalation in therapy such as adding diabetes medication or increasing doses.

Mood disturbance was identified with ICD-10 codes of F32.x, F33.x (major depressive disorder), or F41.1 (generalized anxiety disorder) or patient-reported mood assessments. Assessments include the PHQ-9, a proxy for emotional burden, and the GAD-7 for anxiety. A score greater than or equal to 10 on either indicated mood disturbance.

We observed differences in mean HbA1c and PHQ-9 values across four treatment pathways from Oregon Health & Science University (OHSU) sites. PHQ-9 was selected because it is routinely collected and DD-specific screening measures are underutilized. Patients met the treatment pathway if they had the prescribed treatment within the next year.

- **Mood Treatment** includes patients who received pharmacologic or counseling approaches.
- **Certified Diabetes Care and Education Specialist (CDCES)** includes patients who received at least one session facilitated by a CDCES.
- **Combined Care** included patients who received both treatments. Ordering of treatment was not accounted for here and is an area of future work.
- **Neither treatment** includes patients who were referred to CDCES or prescribed mood disorder medication but did not take either.

## Results

Of 717 patients who had uncontrolled Type 2 DM and mood disturbance, 4.3% received both CDCES and mood disorders treatment; 5.6% received CDCES treatment only; 50.5% received mood disorder treatment only; and 39.6% received no treatment.

Hemoglobin A1c and PHQ-9 outcomes varied meaningfully by pathway, with minimal improvements observed among patients receiving no treatment or mood treatment alone. CDCES-only care was associated with the greatest improvement in A1c, but lower PHQ-9 change.

Treatment initiation rates are low: referral to CDCES occurs in < 6% of patients, and only 25– 50% of adults with DM and depression receive pharmacologic or behavioral treatment. Starting with CDCES-led support can reduce emotional burden, DD, and improve self-management. Combined care demonstrated the most balanced improvements across both outcomes but only 4.3% of patients received both treatment pathways. Increasing the treatment received improves all outcomes.

## Discussion

Clinicians face a decisional dilemma to identify the most appropriate and effective treatment pathway for the growing number of people with DD. Guideline-based care suggests referral to a mental/behavioral health specialist is effective but uptake is low due to lack of access, inconsistent referral pathways, and low patient engagement.

A full randomized control trial is needed to detect a clinically important difference of 0.5% in A1c and a clinically meaningful effect size of BH symptoms. Two important subgroup analyses should be included (1) people on insulin; and (2) with more severe mood disorder symptoms (PHQ-9/GAD-7>15/emotional subscale of DDS > 3.0). Use of technical solutions to increase patient engagement with DM education and BH support is crucial to explore.

**Table 1.** Outcomes by Care Pathway Received at OHSU.

| Patient Group | N (%)<br>Total 717 | Base-line<br>A1c | F/U A1c | Base-line<br>PHQ-9 | F/U PHQ-9 | Mean A1c<br>Change | Mean<br>PHQ-9<br>pts (%)** |
| --- | --- | --- | --- | --- | --- | --- | --- |
| Neither<br>Treatment | 284<br>(39.6%) | 8.6% | 7.9% | 12.2 | 10.0 | -0.64% | -2.2 pts (-18%) |
| Mood<br>Treatment<br>Only | 362<br>(50.5%) | 8.7% | 7.7% | 12.2 | 9.4 | -0.96% | -2.8 pts (-23%) |
| CDCES /<br>Distress<br>Only | 40 (5.6%) | 9.5% | 8.1% | 12 | 11.4 | -1.44% | -0.6 pts (-5%) |
| CDCES +<br>Mood<br>(combine<br>d care) | 31 (4.3%) | 8.8% | 7.6% | 11.9 | 8.8 | -1.2% | -3.1 pts (-26%) |

## Data Availability

Data are not available for this study

## Declarations

### Ethics Approval (IRB)

IRB of Oregon Health & Science University waived ethical approval for this work

### Data Availability

A de-identified data set is available from the authors.

